# Association between TG/HDL-C and depression: Results from the NHANES, 2005-2020

**DOI:** 10.1101/2025.01.12.25320437

**Authors:** Xuemiao Tang, Qiuhua He, Na Yang, Qiang Fu

## Abstract

Lipid profile disturbances are frequently observed in depressive patients. Triglyceride to high-density lipoprotein cholesterol (TG/HDL-C) can be reflective of the level of blood lipids. However, it remains unclear whether higher TG/HDL-C increases the risk of depression. This study aimed to investigate the association between TG/HDL-C and depression. In this cross-sectional study, 19,297 participants were analyzed from the National Health and Nutrition Examination Survey (NHANES) between 2005 and 2020. A weighted multivariable logistic regression model, restricted cubic spline regression model and threshold effect analysis were used to explore the association and dose-response relationship of TG/HDL-C with depression risk in the total population and subgroups. A total of 19,297 participants who had complete data on TG/HDL-C and depression measurement were included in this study (mean age 50 years, 51% female). Participants with depression had higher TG/HDL-C than those without depression. TG/HDL-C was associated with an increased risk of depression after adjusting for all covariates (in model 3: OR = 1.12, 95% CI: 1.03-1.21, *P* = 0.005). Moreover, a nonlinear J-shaped relationship was observed between TG/HDL-C ratio and the risk of depression, with an inflection point of 0.402 by threshold effect analysis. These findings indicate that TG/HDL-C has a J-shaped association with the risk of depression.

## Introduction

Depression, a complex neuropsychiatric condition, is characterized by persistent dysthymia and anhedonia, manifesting as a marked diminution in pleasure or interest in activities of daily living. The estimated prevalence rate of depression is approximately 4.7%, including 5% of adults[1, 2]. Depression can lead to psychosocial dysfunction, worsen the affected person’s life situation, and increase suicides and mortality[3]. According to the Global Burden of Disease Study, depression represents the predominant contributor to disability-adjusted life years among all psychiatric disorders, resulting in a significant economic burden of the healthcare systems[4]. The etiology of depression is multifaceted, including social, biological and psychological factors. Despite advancements in understanding its causes and pathophysiology, the specific risk factors and underlying mechanisms of depression remain incompletely understood[5]. Given the increasing prevalence of depression and its profound impact on both individuals and society, elucidating the multifactorial etiopathogenesis and identifying modifiable risk factors associated with depression remains paramount for developing targeted preventive and therapeutic interventions. By early identifying high-risk individuals and implementing preventive strategies, the heavy burdens experienced by families and communities caused by depression might potentially be alleviated.

Accumulating evidence indicated a close relationship between depression and levels of lipids[6, 7]. Commonly, the principal components of the lipid profile comprise low-density lipoprotein cholesterol (LDL-C), high-density lipoprotein cholesterol (HDL-C), triglycerides (TG), and total cholesterol (TC)[8, 9]. Specifically, previous investigations have demonstrated that individuals with depression presented special pattern of lipids profiles. However, these associations were demonstrated to be inconsistent and even contradictory. For instance, some studies reported decreased TC in depressive individuals[10], while other studies found elevated TC[11] or revealed no specific association[12]. This inconsistency was also showed in HDL-C, LDL-C and TG[6]. Despite heterogeneous findings have been reported in the literatures, abnormal lipids profiles are frequently observed in depressive individuals[13]. In tryptophan hydroxylase-2 knockout mice, the elevated oxidative stress caused alteration of plasma lipid metabolism and thus resulted in 5-hydroxytryptamine deficiency[14]. These findings suggested a biologically plausible link between lipid metabolism and the pathogenesis of depression.

Recent epidemiological evidence has established the triglyceride-to-high-density lipoprotein cholesterol (TG/HDL-C) as an emerging clinically relevant biomarker for metabolic syndrome and cardiovascular disorders[15]. Based on the revealed association between cardiovascular events and the development of depression[16, 17], combined with the biologically plausible link of lipids metabolism to depression, here we hypothesized that an alteration of TG/HDL-C may be independently associated with the incidence of depression. The association between blood TG/HDL-C and the depression risk among U.S. adults was investigated using cross-sectional data from the National Health and Nutrition Examination Survey (NHANES) collected from 2005 through 2020.

## Material and methods

### Study design and subjects

Comprehensive analyses of data extracted from NHANES 2005-2006, 2007-2008, 2009-2010, 2011-2012, 2013-2014, 2015-2016, 2017-2018 and 2019-2020 was conducted, as the participants in these cycles received the depression questionnaire, namely the Patient health Questionnaire (PHQ-9). The NHANES employs a complex, multistage, probability-sampled, cross-sectional design conducted biennially to assess the health and nutritional status of the noninstitutionalized U.S. civilian population through standardized interviewer-administered questionnaires, comprehensive physical examinations, and systematic collection of biological specimens[18]. The complex sampling methodology, standardized research protocols, data collection procedures, and statistical analytical approaches have been extensively documented in peer-reviewed publications and are available online[18]. The study protocol was approved by the National Center for Health Statistics Research Ethics Review Board, and all eligible study participants signed written informed consents before enrollment. Detailed methodological documentation and comprehensive program information for the NHANES are publicly accessible through the NHANES website.

The present investigation included adult participants (age ≥ 18 yr). Participants without blood TG or HDL-C data, or who did not finish the PHQ-9 were excluded. This study was conducted and reported in alignment with the Strengthening the Reporting of Observational Studies in Epidemiology guidelines[19].

### Depression symptoms assessment (outcome)

Depression was assessed using the PHQ-9, a validated nine-item screening instrument that evaluates both the presence and severity of depression. The PHQ-9 is a self-administered diagnostic instrument that consists of nine signs corresponding to the diagnostic criteria for depression as specified in the Diagnostic and Statistical Manual of Mental Disorders, Fourth Edition (DSM-IV)[20]. The PHQ-9 evaluates the frequency and severity of depression experienced by individuals during the preceding 2-week period. Each item is scored on a 4-point ordinal scale (range, 0-3), with the aggregate severity score ranging from 0 to 27 points. In general, a total PHQ-9 score of 10 or higher was considered diagnostic of depression[21], as previous studies demonstrated that a PHQ-9 score of 10 or higher yielded a sensitivity of 85% and a specificity of 89% for the diagnosis of depression[20, 22]. Therefore, all participants were categorized into two groups based on their PHQ-9 scores: those scoring less than 10 points (classified as non-depression) and those scoring 10 points or greater (classified as having depression).

### Laboratory measurements (exposure)

Blood specimens were collected from participants at a mobile examination center (MEC) by NHANES personnel after a minimum 8-hour fasting period. Blood TC, TG and HDL-C concentrations were quantified using direct enzymatic methods. Based on measured values of TG, TC and HDL-C, LDL-C concentrations were calculated by means of the Friedewald formula (LDL-C = TC - HDL-C - [TG/5]) for participants with TG levels below 400 mg per deciliter; for those with higher TG levels, direct LDL-C measurements were performed[23]. The TG/HDL-C (continuous) was calculated as the quotient of serum TG concentration to HDL-C concentration (measured in mg/dL). Detailed information regarding laboratory techniques and quality assurance procedures is available in the Laboratory Procedures Manual of NHANES[18].

### Covariates assessment

Sociodemographic characteristics were assessed through standardized self-administered questionnaires, including age (continuous), gender, race, education attainment and income-to-poverty ratio (IPR) (continuous). The IPR was calculated as the annual total annual household income divided by the federal poverty threshold, which was based on corresponding state and survey year. Participants were stratified into three income categories according to their IPR: high income (IPR of 2 or greater), middle income (IPR of 1 to less than 2), and low income (IPR less than 1)[24]. Height and weight measurements were obtained by certified health professionals using calibrated equipment and standardized protocols at the MEC. Body-mass index (BMI) was calculated as weight in kilograms divided by the square of height in meters. Evaluated comorbidities included diabetes mellitus and hypertension. The diagnosis of diabetes mellitus or hypertension was established based on either physician documentation of the condition or current use of antihypertensive or antihyperglycemic medications. The following variables were also included as covariates: waist circumference in cm (WC) (continuous), smoking status, alcohol intake, LDL-C (continuous) and TC (continuous). Detailed definitions of all covariates were available at NHANES website. Multicollinearity among independent variables was assessed using variance inflation factors (VIF)[25].VIF analysis demonstrated no evidence of significant collinearity among the predictor variables (Supplementary Table 1).

### Statistical analysis

All statistical analyses incorporated the complex multistage probability sampling design of NHANES[26]. Descriptive statistics were presented as follows: continuous variables with normal distribution are presented as means (± standard deviations [SD]), non-normally distributed continuous variables as medians (interquartile range [IQR]), and categorical variables as frequencies (percentages). Between-group comparisons were performed using the weighted Student’s *t* tests, Mann-Whitney *U* tests, and χ^2^ tests, respectively.

In the primary analysis, the association between TG/HDL-C and depression was assessed using multivariable logistic regression models. The TG/HDL-C was put in the model as continuous variable after log transformation due to the skewness of its distribution. The odds ratio (OR) and corresponding 95% confidence interval (CI) were calculated. Three logistic regression models were established: Model 1, no potential confounding factors were adjusted; Model 2, adjusted for sociodemographic characteristics including age, gender, and race; Model 3 was the fully adjusted model, which further adjusted for BMI, WC, education attainment, PIR, hypertension, diabetes mellitus, smoking status, alcohol intake, HDL-C, and TC.

Several sensitivity analyses were conducted. First, the TG/HDL-C was classified into quartiles, and the linear trend across quartiles was evaluated using the median value of each quartile as a continuous variable in the regression model. Second, the potential non-linear association between the TG/HDL-C and depression risk was evaluated using weighted restricted cubic spline (RCS) analyses, with knots placed at the 25th, 50th, and 75th percentiles. These analyses were conducted within the framework of Model 3. Third, to identify potential threshold effects, a two-piecewise linear regression model was fitted after adjustment for all confounding factors. The optimal threshold point was determined by maximizing the log-likelihood function. The presence of a threshold effect was assessed through comparison of a two-piecewise linear model with the single-linear model using likelihood ratio testing.

Subgroup analyses were performed to examine the modification effect of age (age was categorized into three levels, < 40, 40-60 and > 60 yr), gender, race, BMI (BMI was divided into two levels, < 24 and ≥ 24 kg/m^2^), education attainment, PIR, hypertension, diabetes mellitus, smoking status, and alcohol intake on the association between TG/HDL-C and depression. To evaluate potential interaction effects, multiplicative interaction terms were constructed between the TG/HDL-C and prespecified subgroup variables in the multivariable models. For missing values, we generated multiple imputations of the dataset by chained equation with predictive matching algorithms. Five imputed datasets were generated to address potential bias from incomplete observations in the analysis cohort (mice R package, version 4.1.3)[27].

All analyses accounted for complex sampling design of NHANES, including stratification, clustering, and sampling weights[28]. The statistical analyses were executed with R version 4.1.3 (R Foundation for Statistical Computing, Vienna, Austria). The threshold for determining statistical significance employed two-tailed probability assessments with an alpha level set at 0.05.

## Results

### Baseline characteristics of the study population

Of 116,876 participants enrolled in the NHANES from 2005 through 2020, we identified 64,313 adults (≥ 18 yr) who were eligible for this study. We excluded 45,016 participants who had missing data on TG or HDL-C measurement, or who responded “refused” or “don’t know” to items on the PHQ-9. The final analytical cohort comprised 19,297 participants from the NHANES (2005-2020) with complete data on TG/HDL-C and depression assessments. The participants had an average age of 50 years, and 51% were women. The process of participant selection is shown in Fig. 1. Of the 19,297 participants in the analytical cohort, 1,695 (8.8%) met the diagnostic criteria for depression. The proportion of female participants was higher in depressive participants (1,103 [65%] of 1,695 participants) than in non-depressive participants (8,716 [50%] of 17,602 participants). Non-Hispanic White participants constituted the largest racial and ethnic group in both the depression and non-depression cohorts (691 of 1,695 [41%] and 7,412 of 17,602 [42%], respectively). Participants living below the federal poverty threshold (PIR < 1) were more prevalent in the depression group (899 [53%] of 1,695 participants) than in the non-depression group (5,464 [31%] of 17,602 participants). The proportion of participants who were current smokers was higher in depressive participants (35%) compared to non-depressive participants (19%). The proportion of people who had hypertension was higher in depressive participants (53%) than in non-depressive participants (41%). Similarly, the proportion of people who had diabetes mellitus was higher in depressive participants (30%) compared to non-depressive participants (21%). Depressive participants had higher BMI than non-depressive participants. The mean BMI was higher among participants with depression than among those without depression (31.0 ± 8.0 vs. 29.0 ± 7.0 kg/m²). Besides, in comparison to the non-depression group, participants in depression group had lower HDL-C (1.36 ± 0.43 vs. 1.40 ± 0.42 mg/dL) and higher TG (1.58 ± 1.45 vs. 1.39 ± 1.20 mg/dL). Baseline sociodemographic and clinical characteristics of the study cohort are presented in Table 1.

**Fig. 1.**
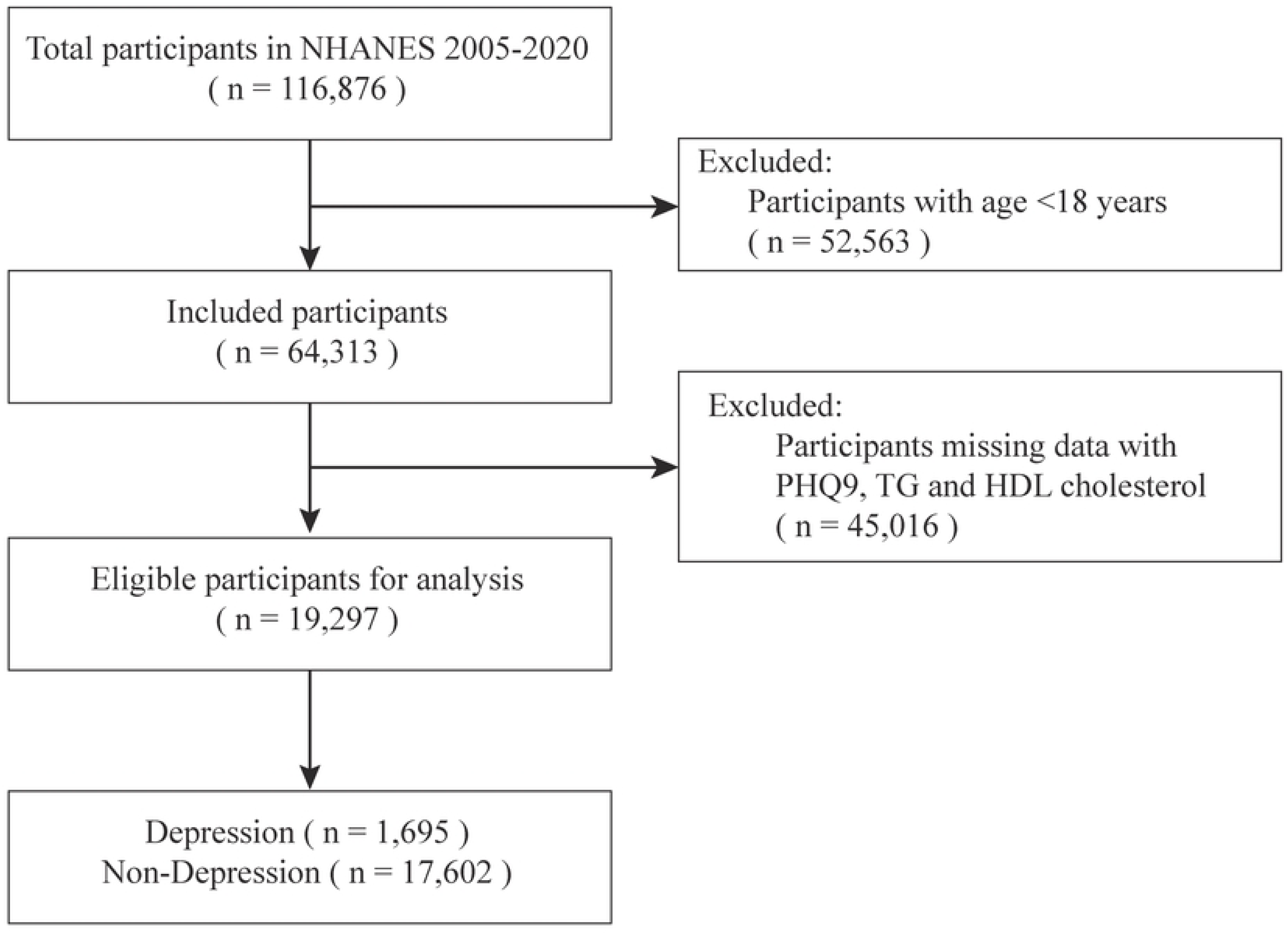
Flowchart of participants’ enrollment process.

**Table 1.** Characteristics of the study population, NHANES 2005-2020.

| Characteristics | Non-depression,<br>N = 17,602 | Depression,<br>N = 1,695 | P value |
| --- | --- | --- | --- |
| **Age, years** | 50 (18) | 50 (16) | 0.4 |
| **Gender** |  |  | <0.001 |
| Female | 8,716 (50%) | 1,103 (65%) |  |
| Male | 8,886 (50%) | 592 (35%) |  |
| **Race or ethnicity** |  |  | <0.001 |
| Mexican American | 2,701 (15%) | 243 (14%) |  |
| Non-Hispanic Black | 3,675 (21%) | 398 (23%) |  |
| Non-Hispanic White | 7,412 (42%) | 691 (41%) |  |
| Other Hispanic | 1,691 (9.6%) | 230 (14%) |  |
| Other, including multiracial | 2,123 (12%) | 133 (7.8%) |  |
| **PIR** |  |  | <0.001 |
| <1 | 5,464 (31%) | 899 (53%) |  |
| ≥1 to <2 | 1,358 (7.7%) | 155 (9.1%) |  |
| ≥2 | 10,780 (61%) | 641 (38%) |  |
| **Education level** |  |  | <0.001 |
| 9-11th grade | 2,324 (13%) | 335 (20%) |  |
| College graduate or above | 4,358 (25%) | 176 (10%) |  |
| High school graduate | 4,055 (23%) | 414 (24%) |  |
| Less than 9th grade | 1,598 (9.1%) | 254 (15%) |  |
| Some college or AA degree | 5,267 (30%) | 516 (30%) |  |
| **Diabetes mellitus** | 3,694 (21%) | 514 (30%) | <0.001 |
| **Hypertension** | 7,227 (41%) | 900 (53%) | <0.001 |
| **Smoking status** |  |  | <0.001 |
| Current | 3,287 (19%) | 589 (35%) |  |
| Former | 4,459 (25%) | 400 (24%) |  |
| Never | 9,856 (56%) | 706 (42%) |  |
| **Alcohol intake** |  |  | 0.001 |
| Current | 12,861 (73%) | 1,174 (69%) |  |
| Former | 2,384 (14%) | 278 (16%) |  |
| Never | 2,357 (13%) | 243 (14%) |  |
| **BMI, kg/m<sup>2</sup>** | 29 (7) | 31 (8) | <0.001 |
| **WC, cm** | 100 (17) | 104 (18) | <0.001 |
| **TG, mg/dL** | 1.39 (1.20) | 1.58 (1.45) | <0.001 |
| **HDL-C, mg/dL** | 1.40 (0.42) | 1.36 (0.43) | <0.001 |
| **TC, mg/dL** | 4.93 (1.08) | 4.99 (1.17) | 0.2 |
| **LDL-C, mg/dL** | 2.90 (0.93) | 2.91 (1.01) | 0.8 |
| **TG/HDL-C** | 0.83 (0.50, 1.39) | 0.98 (0.57, 1.68) | <0.001 |

| Characteristics | Non-depression,<br>N = 17,602 | Depression,<br>N = 1,695 | P value |
| --- | --- | --- | --- |

### Association between TG/HDL-C and depression

Patients with depression demonstrated significantly higher median TG/HDL-C than those without depression (0.98 [0.57-1.68] vs. 0.83 [0.50-1.39]; *P* < 0.001) (presented in Fig. 2).

**Fig. 2.**
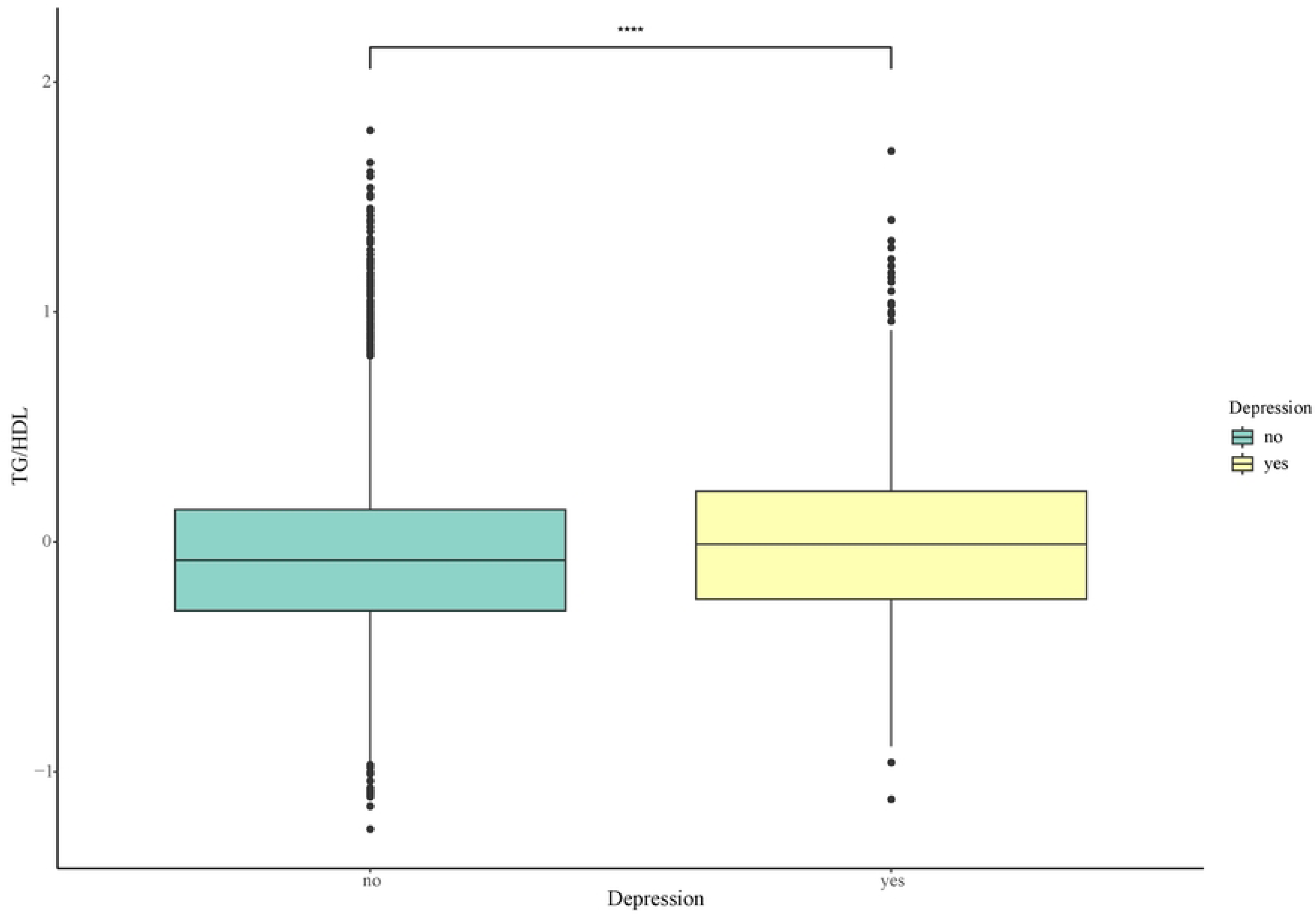
Log10-transformed TG/HDL-C in non-depression group and depression group. “****” indicates *P* <0.0001, as compared with non-depression group.

Multivariable logistic regression analysis revealed that TG/HDL-C was independently associated with depression (presented in Table 2). This association remained significant after adjustment for multiple potential confounders: Model 1 (OR, 1.28; 95% CI, 1.20 to 1.36; *P* < 0.001), Model 2 (1.44; 1.35 to 1.54; *P* < 0.001), and Model 3 (1.12; 1.03 to 1.21; *P* = 0.005).

In the sensitivity analyses, the TG/HDL-C were converted into quartiles: Q1 group (TG/HDL-C ≤ 0.51, n = 4,945), Q2 group (0.52 ≤ TG/HDL-C ≤ 0.84, n = 4,712), Q3 group (0.85 ≤ TG/HDL-C ≤ 1.42, n = 4,867), Q4 group (TG/HDL-C ≥ 1.43, n = 4,773). The risk of depression was significantly higher among participants in the Q4 group than among those in the Q1 group, with consistency across all models (Model 1: 1.61; 1.40 to 1.85; *P* < 0.001; *P* for trend < 0.001; Model 2: 2.04; 1.76 to 2.36; *P* < 0.001; *P* for trend < 0.001; Model 3: 1.22; 1.04 to 1.44; *P* = 0.018; *P* for trend < 0.001). Participants in the Q3 group exhibited a higher incidence of depression compared with those in the Q1 group in Model 1 and Model 2 (Model 1: 1.24; 1.07 to 1.43; *P* = 0.004; *P* for trend < 0.001; Model 2: 1.41; 1.21 to 1.64; *P* < 0.001; *P* for trend < 0.001), but not in Model 3 (1.02; 0.87 to 1.20; *P* = 0.8; *P* for trend < 0.001). There was no significant difference in depression incidence between participants in the Q2 group and those in the Q1 group (*P* > 0.05).

**Table 2.** The association of TG/HDL-C with depression, NHANES 2005-2020.

| Outcomes | Categorical models |  |  |  |  | Continuous models |  |
| --- | --- | --- | --- | --- | --- | --- | --- |
| | Q1 | Q2 | Q3 | Q4 | $P$ for | Log10 TG/HDL-C | $P$ |
| | ($\leq 0.51$) | (0.52-0.84) | (0.85-1.42) | ($\geq 1.43$) | trend | | value |
| Model 1 | 1.00 | 1.02 | 1.24 | 1.61 | $<0.001$ | 1.28 | $<0.001$ |
|  | (ref) | (0.88, 1.19) | (1.07, 1.43) | (1.40, 1.85) |  | (1.20, 1.36) |  |
| Model 2 | 1.00 | 1.09 | 1.41 | 2.04 | $<0.001$ | 1.44 | $<0.001$ |
|  | (ref) | (0.93, 1.27) | (1.21, 1.64) | (1.76, 2.36) |  | (1.35, 1.54) |  |
| Model 3 | 1.00 | 0.91 | 1.02 | 1.22 | $<0.001$ | 1.12 | 0.005 |
|  | (ref) | (0.78, 1.07) | (0.87, 1.20) | (1.04, 1.44) |  | (1.03, 1.21) |  |

Model 1 was not adjusted for covariates.
Model 2 was adjusted for age, gender, race or ethnicity.
Model 3 was adjusted for all covariates including age, gender, race or ethnicity, PIR, education level, alcohol intake, diabetes mellitus, hypertension, smoking status, BMI, WC, TC, LDL-C.

RCS analysis revealed a nonlinear, J-shaped association between the TG/HDL-C and depression after full adjustment for potential confounders (*P* for nonlinearity = 0.008) (Fig. 3), with an inflection point at a log10-transformed TG/HDL-C of −0.395 by threshold effect analysis (TG/HDL-C equal to 0.402) (as presented in Table 3). Therefore, below the threshold ratio of 0.402, there was no significant association between the TG/HDL-C and the risk of depression (OR, 1.13; 95% CI, 0.35 to 3.67; *P* = 0.84). However, above the threshold ratio of 0.402, higher TG/HDL-C was significantly linked to a higher risk of depression. (1.65; 1.29 to 2.10; *P* < 0.001) (Table 3).

**Fig. 3.**
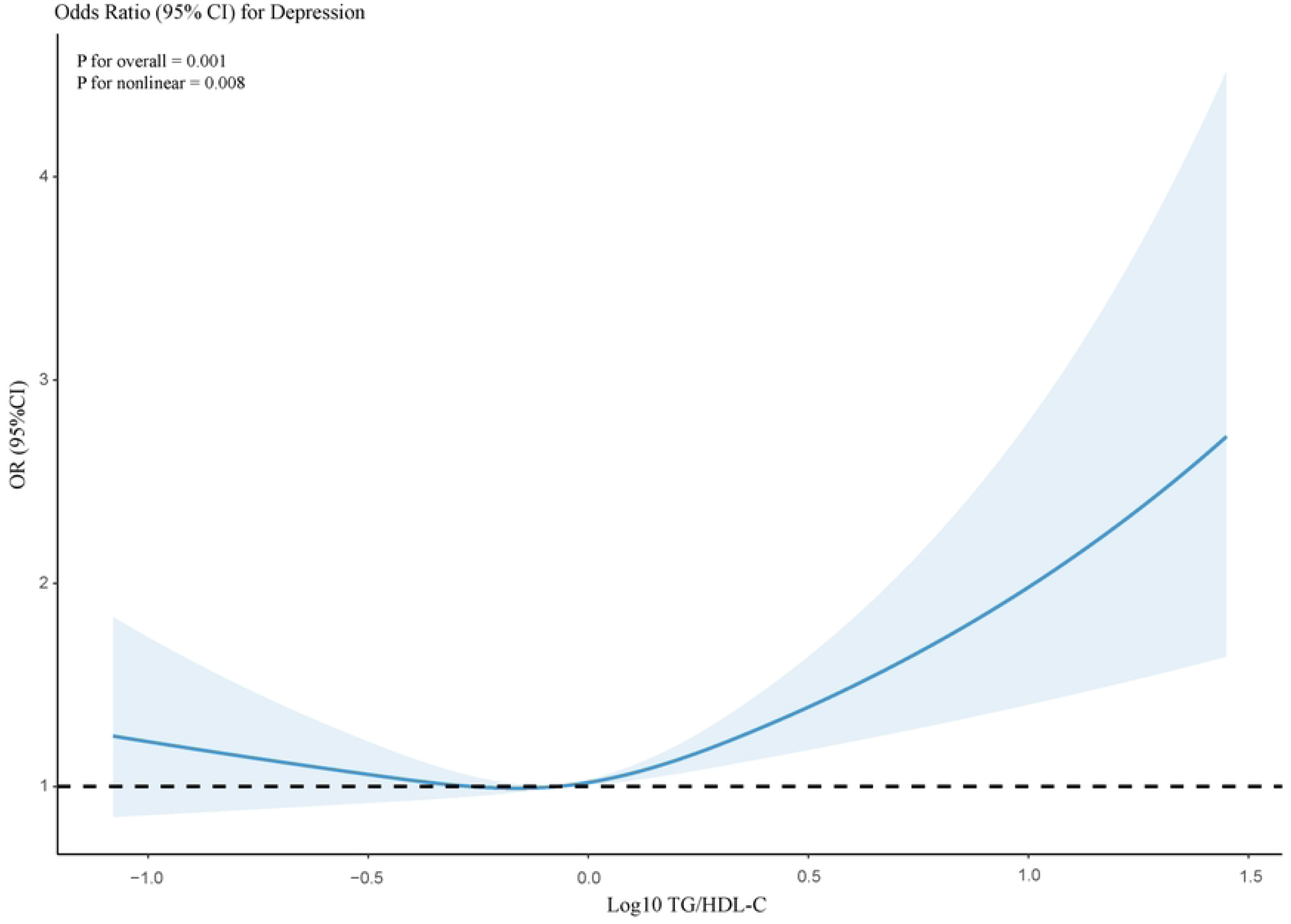
Odds ratio of depression according to log10-transformed TG/HDL-C. The solid line and shadow represented the odds ratio of depression and 95% confidence interval, respectively. All covariates were adjusted in this model.

**Table 3.**
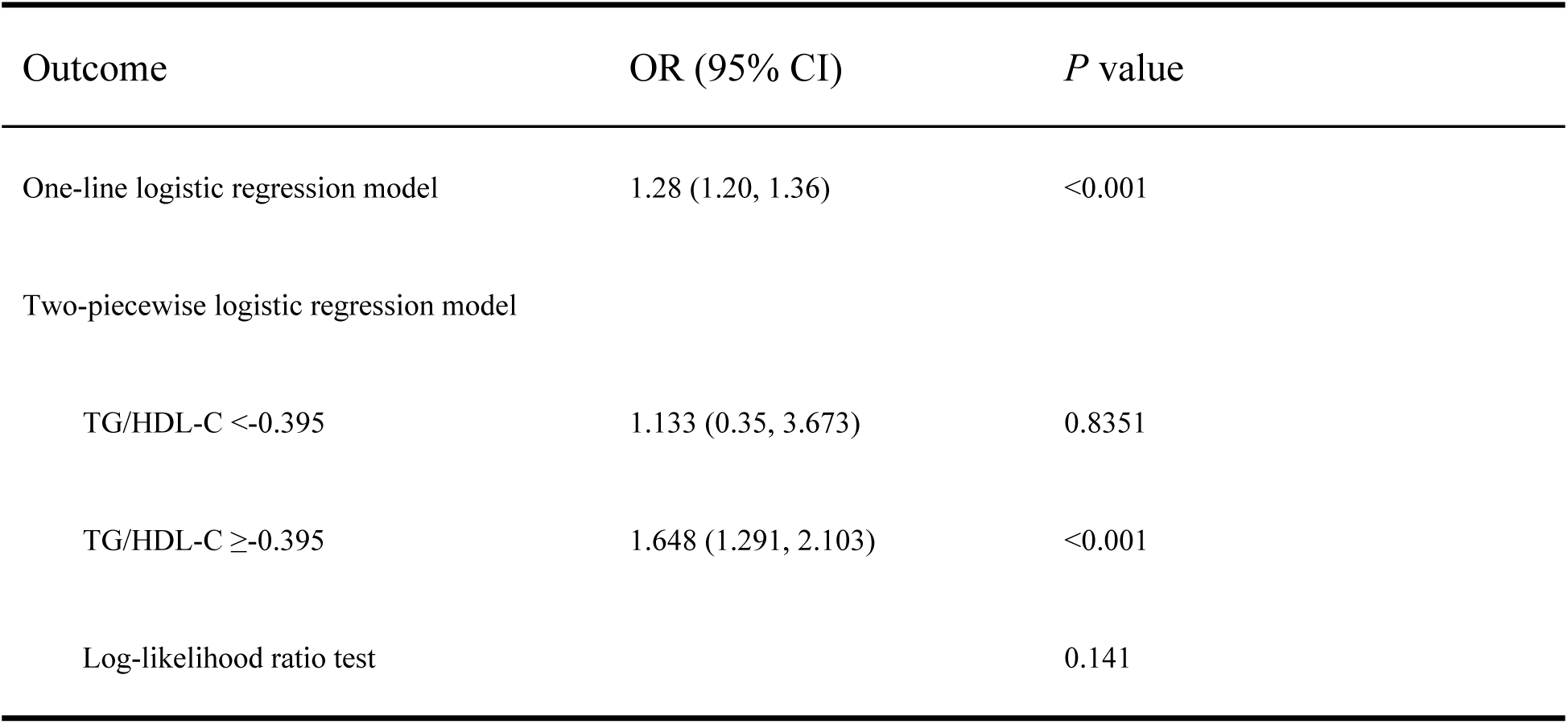
Threshold effect analysis of TG/HDL-C on the risk of depression.

### Subgroup analyses

We performed prespecified subgroup analyses to assess whether the association between TG/HDL-C and the depression risk was consistent across the different subgroups. As presented in Fig. 4, statistically significant interactions were demonstrated between TG/HDL-C and gender, hypertension in relation to depressive symptoms (*P* for interaction = 0.004, 0.017, respectively). Besides, we further constructed the RCS model to explore the association between TG/HDL-C and the risk of depression stratified by gender and hypertension. As presented in Supplementary Fig. 1, female participants and participants with hypertension had higher risk of depression.

**Fig. 4.**
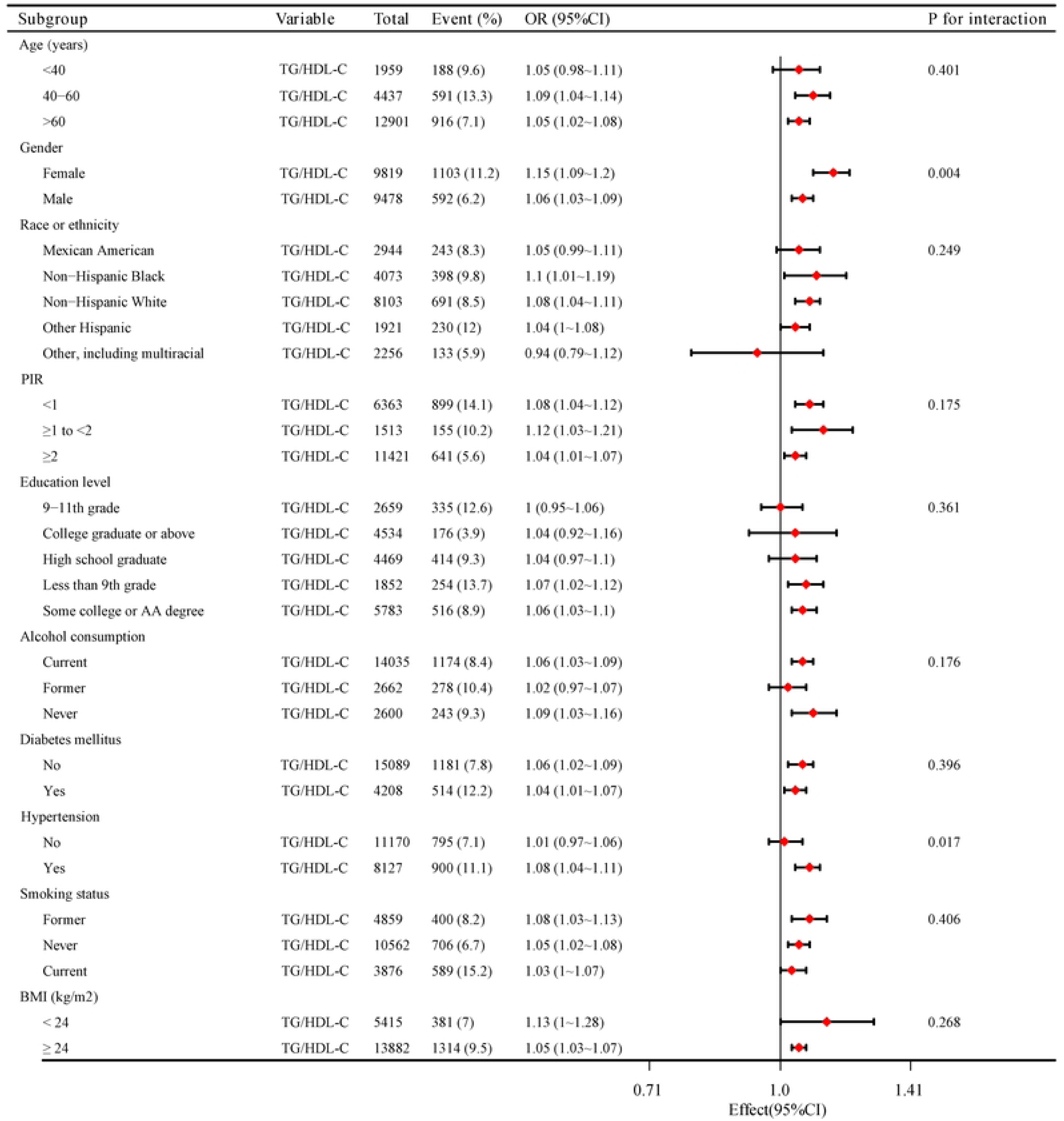
The association between TG/HDL-C and the risk of depression according to different subgroups.

The interaction was not significant after stratification by age, race, PIR, education attainment, BMI, smoking status, alcohol intake and diabetes mellitus. The association between TG/HDL-C and depression remained consistent across different subgroups, with no significant interactions observed (*P* for interaction > 0.05).

## Discussion

Using data from NHANES (2005 through 2020), we examined the association between TG/HDL-C and depression in a cross-sectional cohort of 19,297 participants. TG/HDL-C was significantly higher among subjects with depression than among those without depression. Moreover, we observed a nonlinear, J-shaped association between TG/HDL-C and depression. According to subgroup analysis, significant interaction effects were found between TG/HDL-C and subgroup variables of gender and hypertension. Participants with depression had a higher level of TG/HDL-C, this finding was consistent across different gender, with the association being greater in female participants. It was notable that the association between TG/HDL-C and the depression risk was statistically significant only in participants with hypertension, which was not reported in participants without hypertension.

Depression is a prevalent psychiatric condition worldwide and represents a significant global public health burden, characterized by substantial morbidity and increased risk of suicide-related mortality[29–31]. The clinical manifestations of depression are diverse, and the specific pathogenesis of depression is unclear. Emerging evidence suggests that altered lipid metabolism may contribute to the pathophysiology of mental disorders, including depression, and has been associated with suicidal behavior[32–34]. Alteration in serum lipid profile is associated with the development of depression[35, 36]. It was demonstrated that low serum cholesterol levels can affect neuronal membrane fluidity and lipid-mediated adhesion properties, potentially resulting in diminished serotonergic receptor density and functionality in cerebral tissues[37, 38]. Moreover, low serum cholesterol levels can affect synaptic plasticity, which can further lead to the development of depression. Notably, it appears that there are shared biological pathways and genetic factors that underly both depressive symptoms and abnormal lipid metabolism[39–41]. Furthermore, it has been demonstrated that this genetic effect is especially pronounced for TG and HDL-C[42]. In our study, serum lipid profiles were compared between participants with depressive disorder and without depressive disorder. The findings demonstrated significantly higher TG concentrations and lower HDL-C levels among patients meeting diagnostic criteria for depression. Neither TC nor LDL-C concentrations demonstrated significant between-group differences. These findings were aligned with the findings of a recent meta-analysis, which demonstrated that higher TG concentrations and lower HDL-C levels were significantly associated with depression[36]. These findings suggest that disturbance in serum lipid homeostasis may contribute significantly to the pathogenesis of depressive disorder.

In recent years, the TG/HDL-C has emerged as a marker of considerable clinical interest. Accumulating evidence from clinical investigations suggested that elevated TG/HDL-C may independently predict the risk of several cardiometabolic disorders, including atherosclerotic cardiovascular disease, type 2 diabetes mellitus, metabolic syndrome, hyperuricemia, and chronic kidney disease[43, 44]. To our knowledge, this investigation represents the first systematic examination of the association between TG/HDL-C and depression. The results demonstrated a significant association between an elevated TG/HDL-C and depression risk. After accounting for potential confounding variables, the TG/HDL-C remained significantly elevated in patients with depression compared to those without depression. Moreover, in multivariate analyses, after adjustment for potential confounding variables, we identified a significant nonlinear dose-response association between TG/HDL-C and the risk of depression. When the log10-transformed TG/HDL-C was higher than −0.395 (TG/HDL-C = 0.402), the risk of developing depression increased significantly. These findings suggest that an elevated TG/HDL-C may represent a potential biomarker for early detection of depressive disorders. Therapeutic interventions targeting the normalization of serum TG/HDL-C warrant investigation as a possible strategy for both prevention and amelioration of depressive symptoms. Prospective randomized controlled trials are required to establish causal relationships and assess the clinical relevance of modifying the TG/HDL-C in the management of depression. Statins are one of the most prescribed medications worldwide, which is used for regulating serum lipid levels[45]. It has been reported that stains alone might have a beneficial effect on depression and suicidal behavior[46–49]. Furthermore, recent studies demonstrated that statins had a protective effect against depression when used as an add-on therapy with anti-depressant medications such as selective serotonin reuptake inhibitors (SSRIs)[50, 51]. Emerging evidence, including our findings, suggested that the serum TG/HDL-C may serve as an objective diagnostic biomarker for depression. Furthermore, therapeutic modulation of this ratio could represent a novel preventive strategy to reduce the incidence of depression in clinical practice.

Serum cholesterol such as HDL-C has been implicated in the pathophysiological mechanisms underlying depression[33, 52]. HDL-C is carried by a lipoprotein, HDL[53]. HDL possess antioxidative properties and can inhibit intracellular reactive oxygen species generation[53]. Thus, levels of HDL-C may be expressive of HDL antioxidative activities[54]. A substantial body of evidences have showed elevated oxidative stress in patients with depression, oxidative stress has been established as a pivotal mechanism in the pathophysiology of depression[54, 55]. Hence, downregulation of antioxidative activities due to decreased HDL-C levels may result in the development of depression, HDL-C may work as an antidepressant through antioxidative activities. Hyperactivity of the hypothalamic-pituitary-adrenal (HPA) axis has been consistently demonstrated in patients with depressive disorder[56]. Blood cholesterol and TG increase due to the hyperactivity of HPA axis. Therefore, elevated TG and cholesterol may be found in depressed patients, which is consistent with our results.

Depression is independently associated with increased cardiovascular morbidity and mortality, as demonstrated by longitudinal epidemiological studies[57, 58]. Similarly, perturbations in serum lipid profiles have been well-documented as independent cardiovascular risk factors[59]. Patients exhibiting dyslipidemia demonstrated not only an elevated risk for depression, but also a significantly increased incidence of adverse cardiovascular outcomes. Patients with concurrent depression and dyslipidemia exhibit a significantly higher risk of adverse cardiovascular outcomes, including acute coronary syndrome and congestive heart failure.

Dyslipidemia independently increased the risk of adverse cardiovascular events among patients with depression, suggesting a potential synergistic interaction between these comorbid conditions[60]. Therefore, routine lipids monitoring and lipids level regulating have a protective effect both on depression and cardiovascular events. This large-scale cross-sectional study indicates a clear distinctive TG/HDL ratio between depression group and non-depression group, potentially opening new prevention or treatment methods for depression and its associated cardiovascular comorbidity.

The present investigation has several notable strengths. First, this study represents the first population-based investigation to examine the association between TG/HDL-C and incident depression in a nationally representative sample of U.S. adults. The current investigation included a substantially larger sample size than prior studies examining the relationship between serum lipid parameters and depression[35]. Second, standardized measurement protocols and validated analytical methods were rigorously implemented in accordance with NHANES quality control procedures, which makes our results more credible. Third, multivariate analyses were used to adjust for potential confounding variables, and sensitivity analyses demonstrated consistent findings across predefined subgroups. Finally, the current study sheds new light on the pathogenic mechanism of depression and early intervention for depression in clinical practice. Our findings demonstrated that the serum TG/HDL-C may serve as a clinically relevant biomarker for the diagnosis of depression. Present depression diagnosis mainly relies on patients’ subjective descriptions and spoken information[61]. Thus, misdiagnoses for depressed patients based on such subjective interview are inevitable[62], which can result in confusion in clinical practice. To address the limitations of depression diagnosis, the development of objective methods to identify depressed patients has been justified. In current study, TG/HDL-C may be a blood biomarker for depression in clinical use. Several limitations warrant consideration when interpreting these findings. Firstly, the diagnosis of depression is based on self-reported questionnaires (PHQ-9), a validated screening instrument, rather than structured clinical interviews or diagnostic criteria, which may lead to inaccurate results due to reporting or recall bias. Secondly, there are many unknown and unmeasured confounding factors which we did not take into account. In the present study, we did not consider factors such as marital status, prior medications use and nutritional data, which may have potential effects on depression and lipid parameters and may have partially diluted the effect estimates. Thirdly, the study sample consisted of participants older than 18 years of age. In fact, depression is common in adolescents in recent years, one in seven 10-19-year-olds experiences a depression globally[63]. Consequently, we may miss some important information as we did not include the participants younger than 18 years old, which may bias current results. Finally, a main limitation of this investigation is its cross-sectional design, which inherently precluded the establishment of temporal relationships or causal inference between serum TG/HDL-C and incident depression. Further investigation of the relationship between TG/HDL-C and the development of depression through longitudinal cohort studies would provide more definitive evidence of causality.

## Conclusion

In conclusion, these findings demonstrated a significant association between elevated TG/HDL-C and depression. We observed a J-shaped association between TG/HDL-C and incident depression. When TG/HDL-C is higher than 0.402, the risk of depression increases significantly, indicating that TG/HDL-C may serve as a biomarker for early identification for depression. These findings suggest that routine TG/HDL-C monitoring and interventions should be conducted in the early-stage of depression to decrease the risk of cardiovascular morbidity. Further prospective longitudinal investigations and randomized controlled trials are warranted to elucidate the potential causal relationship between TG/HDL-C and depression.

## Author statement

### Authorship contributions

Qiang Fu conceived, designed and supervised the study; Xuemiao Tang and Qiuhua He analyzed data and prepared results; Xuemiao Tang wrote the paper with input from all authors; Qiang Fu revised the paper; Xuemiao Tang and Na Yang were involved in data acquisition. All authors finally approved the paper.

### Data availability statement

All data that support the findings of this study can be found on the NHANES website.

## Acknowledgements

The authors thank investigators and participants of the National Health and Nutrition Examination Study.

